# Community Consultation and Public Disclosure for the Randomized Trial of Sedative Choice for Intubation

**DOI:** 10.1101/2025.09.26.25336736

**Authors:** Ariel A. Lewis, Tiffany L. Israel, Kevin P. Seitz, Brian E. Driver, Kevin W. Gibbs, Adit A Ginde, Stacy A. Trent, Derek W. Russell, Matthew E. Prekker, Aaron E. Robinson, Jessica A. Palakshappa, John P. Gaillard, L. Jane Stewart, Logan L. Beach, Bradley D. Lloyd, Stephanie C. DeMasi, Margaret A. Hays, Cori Withers, Amy E. Sullivan, Carolynn Lyle, Micah R. Whitson, Barbara Gould, Todd W. Rice, Wesley H. Self, Jin H. Han, Matthew W. Semler, Jonathan D. Casey, the RSI investigators and the Pragmatic Critical Care Research Group

## Abstract

**Background:** Randomized trials evaluating emergency treatments may be conducted with Exception from Informed Consent (EFIC) when prospective informed consent is infeasible. In EFIC trials, a period of community consultation and public disclosure precedes initiation of enrollment. The Randomized Trial of Sedative Choice for Intubation (RSI) is a 2,364-patient randomized trial being conducted with EFIC in 14 emergency departments and intensive care units across the United States. This manuscript reports the approach to community consultation and public disclosure in the RSI trial.

**Methods:** Community consultation and public disclosure were conducted locally in each of the 5 regions of enrolling sites. The coordinating center provided sites with templates and access to an engagement coordinator to assist with developing and executing site plans.

**Results:** Community consultation and public disclosure occurred at the coordinating center from February 2021-January 2022 and in the regions of the five additional enrolling sites from September 2023-June 2024. Community consultation included in-person surveys with 789 patients or family members in emergency department or intensive care unit waiting rooms and invitation of more than 200 local groups to town halls or community engagement studios. Public disclosure included (i) social media advertisements viewed more than 1.2 million times, (ii) a trial website with more than 16,000 unique visitors, (iii) informational flyers in hospitals and public settings, and (iv) featured information in traditional media. Completing local community consultation and public disclosure required an average of 139 hours of research personnel time and approximately $18,822 at each site, in addition to coordinating center effort and costs.

**Conclusions:** Pre-trial community consultation activities in the RSI trial engaged over 1,000 patients, families, and community members and public disclosure reached over 1.2 million community members. While the total cost and duration of activities at sites were substantially lower than reported in prior EFIC trials, these costs remained significant.

## INTRODUCTION

In response to the ethical dilemma posed by emergency care research in which prospective written informed consent is infeasible and the research is greater than minimal risk, the U.S. Food and Drug Administration (FDA) created the Exception from Informed Consent (EFIC) process in 1996. To qualify for EFIC, a study must focus on a life-threatening medical condition for which available treatments are unproven or unsatisfactory and evaluate an intervention that must be delivered within a time window that is too short for prospective, written informed consent to occur. Studies conducted with EFIC are required to include additional patient protections and mechanisms of addressing ethical principles that would normally be addressed by prospective informed consent. Two important components of studies conducted with EFIC are community consultation and public disclosure.

Community consultation occurs during study design and is a process of engaging representatives of the communities from which patients will be recruited for the study. Community consultation includes bidirectional communication between community members and the research team through mechanisms like town hall meetings. It is intended to show respect for persons while also providing an opportunity for communities affected by the research to give meaningful input to investigators on the design of the study and to the institutional review board (IRB) on the acceptability of the study. Community consultation, therefore, is deep engagement with a narrow portion of the public. Public disclosure, by comparison, is intended to disseminate information about the research to as large a population as possible through channels like websites or advertisements. The primary goal of public disclosure is transparency, and communication is generally one-way from researchers to the public.(1) In addition to the pre-trial public disclosure activities, EFIC trials are also required to disseminate trial results through post-trial public disclosure.

Federal regulations provide limited detail regarding how to conduct community consultation and public disclosure for a trial conducted with EFIC.(2) In the 20 years following the release of EFIC regulations, only 41 randomized trials were registered with the FDA with a plan to use EFIC (approximately 2 trials per year).(3) Few of these trials have published detailed information on the design and execution of their community consultation and public disclosure. To add to the available literature on approaches to community consultation and public disclosure in trials conducted with EFIC, this manuscript describes the community consultation and public disclosure activities for the Randomized Trial of Sedative Choice for Intubation (RSI) (NCT05277896), a randomized trial comparing ketamine versus etomidate for emergency tracheal intubation that is being conducted with EFIC.(4) This is intended to provide an example of a relatively cost-efficient, federated approach to community consultation and public disclosure that attempts to match the scope, nature, and content of community engagement to the risks and burdens of the research.

## METHODS

### The RSI trial

The RSI trial is an ongoing multicenter, pragmatic, randomized trial comparing the use of ketamine versus etomidate for induction of anesthesia during emergency tracheal intubation of critically ill adults in an emergency department or intensive care unit. The RSI trial is being conducted by the Pragmatic Critical Care Research Group (PCCRG), a network of 25 health systems across the U.S. that conducts comparative effectiveness trials focused on acute care.(5) The RSI trial is designed to enroll 2,364 patients from 14 emergency departments or intensive care units at six hospitals across five regions of the U.S. (Table 1). Enrollment began in April of 2022 and is expected to be completed in August of 2025. Enrollment of the first 464 patients at Vanderbilt University Medical Center was funded by the NIH (K23HL153584) and enrollment of the final 1,900 patients at 6 sites in 5 regions across the U.S. is funded by the Patient-Centered Outcomes Research Institute (PCORI) (BPS-2022C3-30021).

**Table 1.** Hospitals that are enrolling patients in the RSI trial.

| Hospital | Location | Description |
| --- | --- | --- |
| Vanderbilt University Medical Center | Nashville, TN | Tertiary care hospital that serves patients with complex medical illnesses and patients from rural communities. |
| Hennepin County Medical Center | Minneapolis, Minnesota | Public teaching hospital and Minnesota's largest safety-net hospital. |
| University of Alabama – Birmingham (UAB) | Birmingham, Alabama | Public teaching hospital and Alabama's largest hospital; serves rural communities state-wide. |
| University of Colorado<br>-UCHealth University of Colorado Hospital | Aurora, Colorado | Large tertiary care hospital that is part of the largest health system in Colorado. |
| -Denver Health Medical Center Campus | Denver, Colorado | Urban safety net hospital in downtown Denver. |
| Atrium Health – Wake Forest Baptist | Winston-Salem, North Carolina | Large tertiary care hospital that is part of the largest health system in North Carolina. |

### Development of the Plan for Community Consultation and Public Disclosure

Development of the plan for community consultation and public disclosure for the RSI trial began in 2020. The initial draft of the plan was informed by two community engagement studios, one with patients and community members and one with clinicians. Community engagement studios are an engagement approach developed by the Meharry-Vanderbilt Community-Engaged Research Core in which a unique panel of 8-12 patients, community members, or clinicians (selected for their firsthand knowledge of a particular condition) serve as paid experts during a 2-hour face-to-face session with researchers.(6) In the initial community engagement studios for the RSI trial, patients recommended (i) preferentially using social media rather than traditional media formats (e.g., television and radio), (ii) collaborating with local clinics including federally qualified health centers and faith-based clinics, and (iii) leveraging local and national patient and caregiver support networks.

In consultation with the single IRB overseeing the trial, personnel at the coordinating center developed a community consultation and public disclosure template that provided sites with a menu of (i) required and (ii) optional activities (Table 2). This approach was intended to provide structure to the community consultation and public disclosure processes while allowing sites to tailor their activities to the unique context of their local communities.(7) Required community consultation activities included face-to-face engagement with patients and community members using structured surveys to solicit input on the acceptability of the proposed RSI trial, which were modeled on the surveys used in prior EFIC trials.(8–11) Required public disclosure activities included developing and maintaining a website for the trial that provided lay language summaries of the trial design, how patients would be enrolled without the ability to provide prospective informed consent, and how to obtain an “opt-out” medical bracelet alert from trial personnel.

**Table 2.**
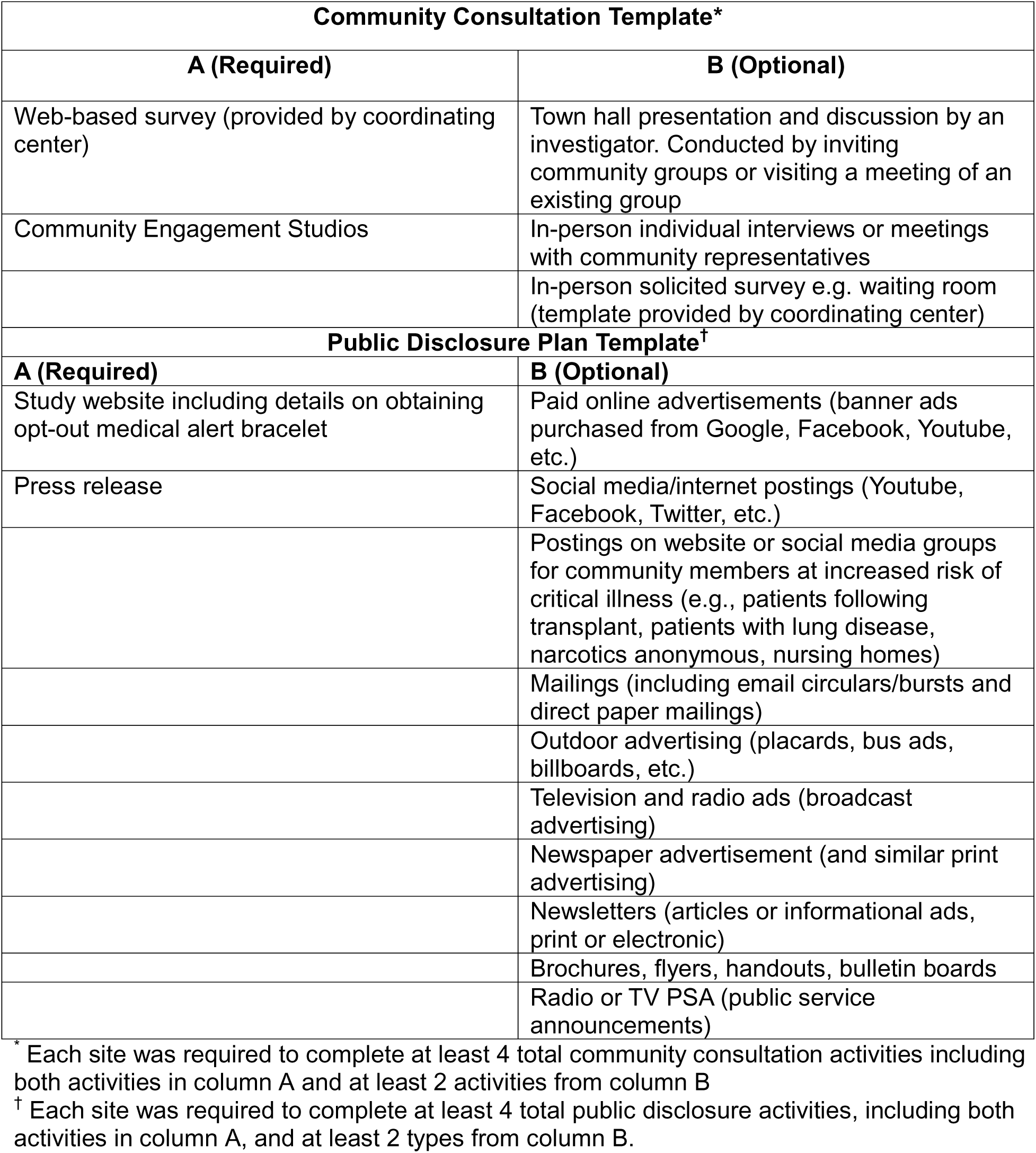
Community Consultation and Public Disclosure Templates.

### Development of Site-Specific Plans

Upon funding of the multi-center portion of the trial in 2023, development of site-specific plans for community consultation and public disclosure at each participating site began. Because sites had varying levels of experience with patient and community engagement, the coordinating center staff included an expert in stakeholder engagement who provided assistance to sites in 1) developing site-specific community consultation and public disclosure plans, 2) developing recruitment materials for community consultation activities, 3) identifying community members and groups to invite to town hall meetings and community engagement studios, 4) coaching site investigators in presentation and communication strategies that should be used when presenting potentially sensitive topics during meetings with community members, and 5) collating community feedback for reporting to local and central institutional review boards.

Approved plans set a minimum bar for planned activities while allowing flexibility for sites to conduct additional activities in response to feedback received from the community during the execution of the planned activities. The coordinating center provided sites with a structured template to track and summarize their completed community consultation and public disclosure activities.

## RESULTS

Vanderbilt University Medical Center, which serves as both the coordinating center and an enrolling site for the RSI trial, completed its community consultation and public disclosure activities over a 12-month period from February 2021 to January 2022. Community consultation and public disclosure activities at the five remaining sites occurred between September 2023 and June 2024. The activities are summarized below and full details are available on FDA public docket 95S-0158.(12)

### Community Consultation Results

Community consultation activities resulted in rich quantitative and qualitative data from a diverse group of community members across the 6 trial sites. A total of 789 English-speaking or Spanish-speaking patients or family members in emergency department or intensive care unit waiting rooms completed face-to-face surveys with trial personnel. Demographics of respondents are shown in Table 3. Approximately 96% of survey respondents agreed with the importance of the RSI trial and 92% agreed with or were indifferent to the conduct of the study without prospective informed consent. Detailed results of the surveys will be reported separately.

**Table 3.** Characteristics of the patients or family members who responded to in-person surveys in ED or ICU waiting rooms.

| **Characteristic** | **Survey Cohort**<br>**(N=789)** |
| --- | --- |
| **Age, median (IQR) – yr** | 44 (28, 60) (N=734) |
| **Female sex – no. (%)** | 427/781 (54.7) |
| **Race and ethnicity – no. (%)** |  |
| Non-Hispanic Black | 199 (25.2) |
| Non-Hispanic White | 364 (46.1) |
| Hispanic | 147 (18.6) |
| Other | 54 (6.8) |
| Prefer not to answer or missing | 25 (3.2) |
| **Highest Level of Education – no. (%)** |  |
| Less than 9<sup>th</sup> Grade | 14/781 (1.8) |
| 9-12<sup>th</sup> grade, no diploma | 71/781 (9.1) |
| High school graduate | 179/781 (22.9) |
| Technical or Vocational Degree | 77/781 (9.9) |
| Some college credit, no degree | 164/781 (21.0) |
| Bachelor's Degree | 152/781 (19.5) |
| Graduate or Professional Degree | 94/781 (12.0) |
| Prefer not to answer | 30/781 (3.8) |
| **Place of Residence– no. (%)** |  |
| Urban | 293/780 (37.6) |
| Suburban | 283/780 (36.3) |
| Rural | 123/780 (15.8) |
| Homeless | 15/780 (1.9) |
| Other | 27/780 (3.5) |
| Prefer not to answer | 39/780 (5.0) |
| **Household Income– no. (%)** |  |
| < \$10,000 | 81/782 (10.4) |
| \$10,000-\$25,000 | 99/782 (12.7) |
| \$26,000-\$50,000 | 131/782 (16.8) |
| \$51,000-\$75,000 | 131/782 (16.8) |
| > \$75,000 | 184/782 (23.5) |
| Prefer not to answer | 156/782 (20.0) |

More than 200 local groups were invited to attend community engagement studios or town halls across the 6 trial sites in 5 regions. Invited groups included faith-based, government, first responder, and disease-specific patient and family support groups (such as support groups for patients with lung disease, hypertension, or stroke). A total of 9 community engagement studios were held across 5 regions, with 82 attendees. These sessions were transcribed by site personnel. Feedback on the trial was generally positive overall and was used to revise patient-facing documents, revise messaging about the trial to participants and families, and inform the way study coordinators were trained to approach and interact with participants and their families. At a community engagement studio dedicated to soliciting input specifically from Black and African American community members, participants highlighted that, given the history of scientific research misconduct involving Black and African American communities, members of the research team should deliberately and actively work to build trust and maintain transparency with members of these communities.

Nine town hall meetings were conducted across 5 regions and included both in-person and online engagement, with a total of 156 attendees. These sessions were transcribed by site personnel. At one site, the patient investigator for the site (a survivor of emergency tracheal intubation who serves on the Steering Committee for the RSI trial) attended the town hall to help community members understand the experience of critical illness and the perspective of a survivor. Attendees of the town halls expressed broad support for the trial at all sites. Attendees also emphasized the importance of timely notification of enrollment in the trial and expressed a desire that more research teams would disseminate information about ongoing research activities within their communities. Selected quotes from the community engagement studios and town halls are available in Table 4. In summary, a majority of consulted community members across the five regions found the RSI trial to be addressing an important research question and acceptable to conduct with EFIC.

**Table 4.**
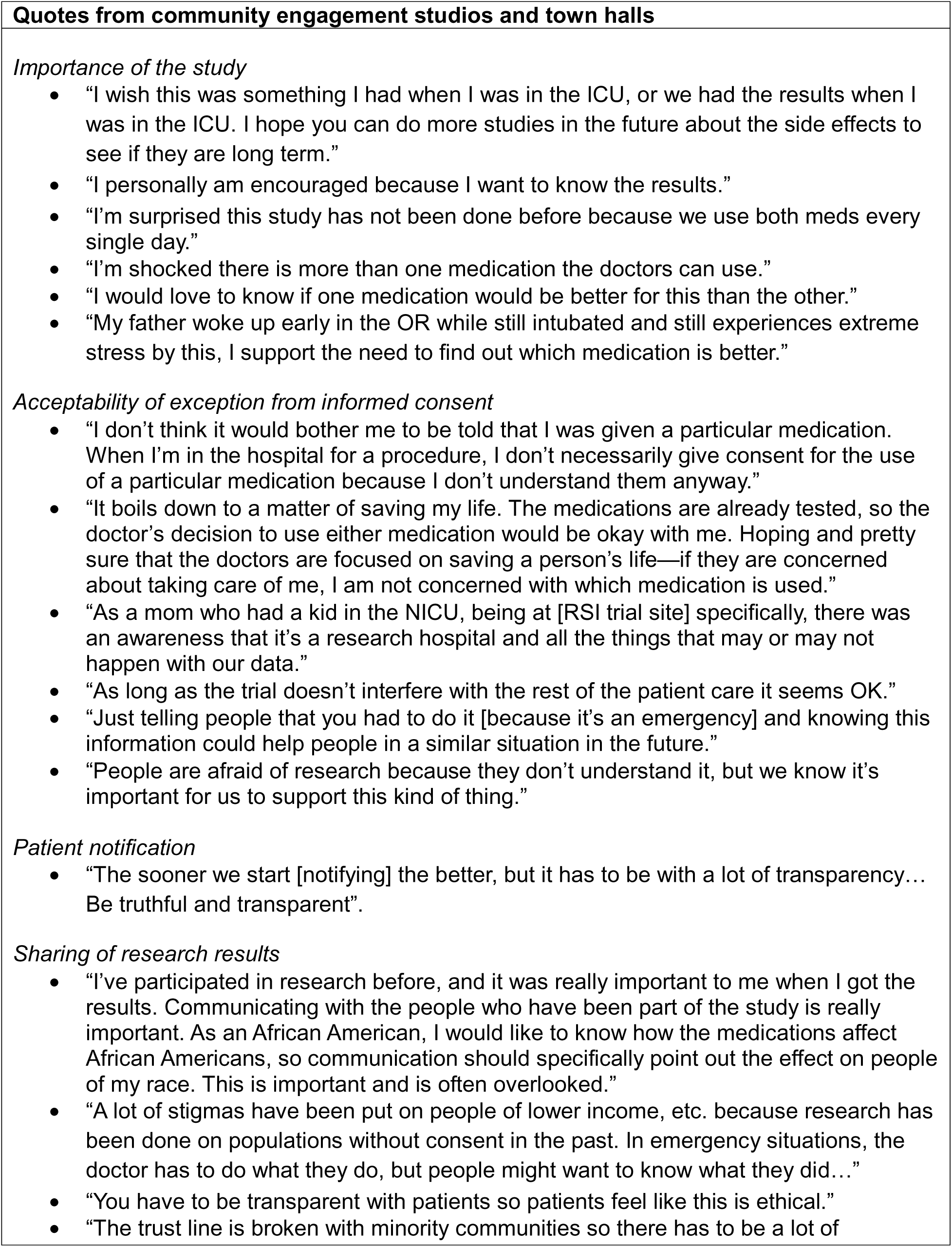
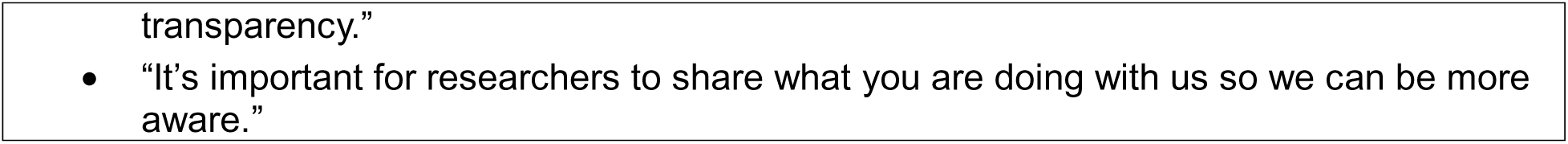
Selected Quotes from the Community Consultation Process.

| Quotes from community engagement studios and town halls |
| --- |
| *Importance of the study*<br>• “I wish this was something I had when I was in the ICU, or we had the results when I was in the ICU. I hope you can do more studies in the future about the side effects to see if they are long term.”<br>• “I personally am encouraged because I want to know the results.”<br>• “I’m surprised this study has not been done before because we use both meds every single day.”<br>• “I’m shocked there is more than one medication the doctors can use.”<br>• “I would love to know if one medication would be better for this than the other.”<br>• “My father woke up early in the OR while still intubated and still experiences extreme stress by this, I support the need to find out which medication is better.”<br>*Acceptability of exception from informed consent*<br>• “I don’t think it would bother me to be told that I was given a particular medication. When I’m in the hospital for a procedure, I don’t necessarily give consent for the use of a particular medication because I don’t understand them anyway.”<br>• “It boils down to a matter of saving my life. The medications are already tested, so the doctor’s decision to use either medication would be okay with me. Hoping and pretty sure that the doctors are focused on saving a person’s life—if they are concerned about taking care of me, I am not concerned with which medication is used.”<br>• “As a mom who had a kid in the NICU, being at [RSI trial site] specifically, there was an awareness that it’s a research hospital and all the things that may or may not happen with our data.”<br>• “As long as the trial doesn’t interfere with the rest of the patient care it seems OK.”<br>• “Just telling people that you had to do it [because it’s an emergency] and knowing this information could help people in a similar situation in the future.”<br>• “People are afraid of research because they don’t understand it, but we know it’s important for us to support this kind of thing.”<br>*Patient notification*<br>• “The sooner we start [notifying] the better, but it has to be with a lot of transparency... Be truthful and transparent”.<br>*Sharing of research results*<br>• “I’ve participated in research before, and it was really important to me when I got the results. Communicating with the people who have been part of the study is really important. As an African American, I would like to know how the medications affect African Americans, so communication should specifically point out the effect on people of my race. This is important and is often overlooked.”<br>• “A lot of stigmas have been put on people of lower income, etc. because research has been done on populations without consent in the past. In emergency situations, the doctor has to do what they do, but people might want to know what they did...”<br>• “You have to be transparent with patients so patients feel like this is ethical.”<br>• “The trust line is broken with minority communities so there has to be a lot of |

### Public Disclosure Results

All six sites used paid social media advertisements on Facebook (Meta) or Instagram to target information about the RSI trial to the geographic catchment area of the participating hospital. All sites posted advertisements in English; two sites also posted advertisements in Spanish. Three sites posted information about the study on social media pages on Linkedin, Twitter (X), Facebook (Meta), or NextDoor. Sites varied in terms of the institutional requirements for marketing or press office involvement in finalizing the social media advertisements from the template provided. Social media posts and advertisements included details of the study, including the fact that patients would be enrolled without informed consent. Posts and advertisements also included a link to the overall study website and a method of requesting opt-out bracelets. The cost of the social media advertising ranged from $750-$2250 across the sites. The cumulative cost of social media advertising across all trial sites was $8,291. Metrics and costs for social media advertisements are shown in Table 5.

**Table 5.** Details of Social Media Campaign used for Public Disclosure.

| **Trial site** | **Social media used for advertisement (length posted)** | **Metrics** | **Cost** |
| --- | --- | --- | --- |
| Site 1 | Facebook (2 months) | Viewed by 176,100 people<br>8,000 people clicked the link | \$2,000 |
| Site 2 | Facebook (6 weeks) | Viewed by 121,752 people<br>584 people interacted with the ad<br>532 people clicked the link | \$1000 |
| Site 3 | Facebook and Instagram (8 days) | English<br>Viewed by 194,389 people<br>144 people interacted with the ad<br>136 people clicked the link<br>Spanish<br>Viewed by 106,505 people<br>79 people interacted with the ad<br>73 people clicked the link | \$800 |
| Site 4 | Facebook and Instagram (1 month) | English and Spanish<br>Viewed by 113,256 people<br>898 people clicked the link | \$1491 |
| Site 5 | Facebook and Instagram (1 month) | Viewed by 45,769 people<br>955 people interacted with the ad<br>875 people clicked the link | \$750 |
| Site 6 | Facebook (2 weeks) | Viewed by 454,158 people | \$2250 |
| **Total** | **---** | **1,211,929 views** | **\$8,291** |

Flyers presenting information about the RSI trial were posted in English and Spanish by five sites in emergency department, critical care, and community clinic settings. Two sites also posted flyers in public spaces, such as local libraries and recreation centers, and mailed flyers to local student and community groups.

Press releases were issued by each of the six hospitals, resulting in two local newspaper articles at one site and one local news television segment at another. One site ran an advertisement about the study in two local print newspapers with an additional advertisement in the online version of one of the newspapers. The study website was available throughout the public disclosure process. During the 6-month period of the most intense public disclosure activities, from January to June 2024, the RSI trial webpage had 16,635 unique visitors and 35,557 page visits.

### Effort and Cost to Conduct Community Consultation and Public Disclosure

The time spent by research teams and principal investigators for community consultation varied considerably across the six sites. Based on self-reported metrics, research staff at sites spent an average of 30 hours (range, 15-60 hours) conducting in-person surveys, an average of 5 hours (range, 0-15 hours) coordinating and hosting community engagement studios, and an average of 22 hours (range 0-110 hours) preparing for and conducting town halls. Site principal investigators spent an average of 2 hours conducting in-person surveys (range 0-10 hours), an average of 25 hours (range, 3-120 hours) coordinating and hosting community engagement studios and an average of 25 hours (range, 6-55 hours) coordinating and hosting town halls.

Cumulatively at all sites, we estimate that approximately 341 hours of research staff time and 310 hours of site principal investigator time were dedicated to completing community consultation. Considering base salary, funder salary caps, fringe benefits, and the 40% indirect rate set by the funder, the average hourly rate for a research coordinator was approximately $80 per hour and the average hourly rate for a physician site investigator was approximately $180 per hour. Using these estimates, the total cost of completing community consultation was $83,080 (approximately $13,847 per site).

The time spent by research teams for public disclosure activities was more consistent across sites than the time spent on community consultation. Based on self-reported metrics, research staff spent an average of 8 hours (range, 0-30 hours) finalizing social media posts and advertisements, an average of 8 hours (range, 2-20 hours) posting flyers, and an average of 2 hours (range,0-10 hours) finalizing press releases. Site principal investigators spent an average of 7 hours (range, 3-10 hours) finalizing social media posts and advertisements, an average of 4 hours (range 2-10 hours) finalizing press releases, and a cumulative total of 4 hours preparing for and conducting a TV segment. Cumulatively, the time spent on public disclosure across the sites for the trial was estimated to include 112 hours of study coordinator time and 70 hours of site principal investigator time. The estimated total personnel cost of completing public disclosure was approximately $21,560. Combining the costs of social media advertising and the time spent by research coordinators and physician site investigators, the estimated total cost of performing public disclosure at all sites was $29,851 (approximately $4,975 per site).

In summary, the estimated total costs of completing community consultation and public disclosure prior to beginning enrollment in the RSI trial was approximately $112,931 (approximately $18,822 per site). These estimates do not include the time spent by coordinating center staff to develop the resources needed for public disclosure or the time spent by coordinating center staff assisting sites with development and execution of their plans, each of which were significant.

### Beyond Community Consultation and Public Disclosure

While the regulations for EFIC require pre-trial community consultation and public disclosure and post-trial public disclosure of trial results, they do not require or recommend patient or community involvement during the conduct of the trial. The RSI trial, however, opted to engage patients, families, and community members at all stages of the research, using a previously published multilevel approach to patient and community engagement (Figure 1).(13)

**Figure 1.**
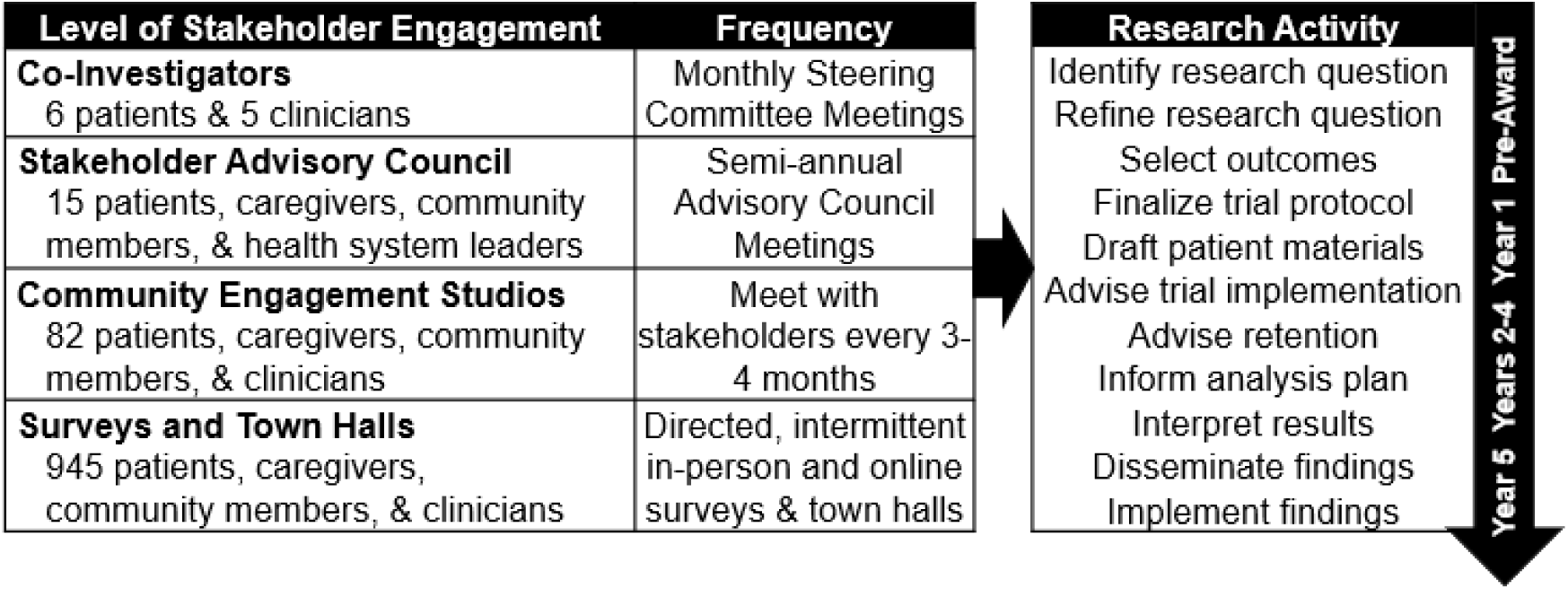
Multilevel approach to stakeholder engagement. Reprinted from DeMasi SC, Imhoff B, Lewis AA, Seitz KP, Driver BE, Gibbs KW, et al. Protocol and Statistical Analysis Plan for a Multicenter Randomized Trial of Ketamine vs Etomidate for Emergency Tracheal Intubation. CHEST Crit Care [Internet]. 2025 Sep;3(3). Available from: https://www.chestcc.org/article/S2949-7884(25)00050-4/fulltext, with permission from Elsevier.

First, trial leadership includes 6 patient investigators, one patient from a patient advocacy group focused on survivors of acute respiratory distress syndrome and one patient from each geographic area in which the trial is enrolling. Each patient investigator participates as full voting members of the Steering Committee and provides input into all major trial decisions. Second, the PCORI-funded Science, Technology and Research partnership (STAR) Clinical Research Network (CRN) provides oversight of the trial through their Stakeholder Advisory Council. The council consists of patients, community members, and clinicians who review research studies to evaluate their person-centeredness and provide advice regarding additional opportunities to improve trial conduct.(14) The STAR-CRN Stakeholder Advisory Council has provided input and feedback on the RSI trial twice yearly. Third, when the trial encounters an issue that would benefit from broader input from patients or community members with lived experience in emergency tracheal intubation (e.g., development of consent forms, amount of financial compensation for participants), trial leadership use the ResearchMatch Expert Advice Tool(15) to survey a geographically representative sample of community members with the relevant prior experience who have volunteered to be available for research studies. Fourth, when the RSI trial needs deep input into a specific study decision (e.g., design of the study’s newsletter for participants), trial leadership convenes a community engagement studio. Studios for the RSI trial have occurred thrice yearly and have addressed topics such as best practices for notification of enrollment, best practice for approaching for consent, and identification of subgroups of interest for subgroup analyses. All of the engagement activities are overseen by the trial engagement coordinator, an expert with more than 20 years of experience leading community engagement in research who provides critical input into early drafts of patient-facing documents, identifies patients and community members for inclusion on the study steering committee and community engagement groups, and leads training and engagement of patient investigators in trial conduct.

## DISCUSSION

Community consultation and public disclosure have been envisioned as methods of addressing the ethical principles of consent in studies in which prospective informed consent is infeasible. Although the regulations outlining the requirements for community consultation and public disclosure in trials conducted under EFIC were published nearly 30 years ago, uncertainty remains regarding the optimal methods and approaches for accomplishing these activities.(1,3,8,10,16,17)

Few studies have reported the full details of their community consultation and public disclosure activities or the associated financial cost, personnel time cost, and duration to complete. Available publications of such metrics for community consultation and public disclosure in a EFIC trials have estimated a cost of $50,000 to $60,000 per site and up to 3 years to complete.(18,19) By contrast, the estimated total fixed and personnel cost to complete community consultation and public disclosure in the RSI trial was approximately $112,931 overall (approximately $18,822 per site), excluding time spent by coordinating center staff to develop the resources and templates or assist sites with development and execution of their plans. Compared to prior EFIC trials, personnel costs were similar, but the fixed costs of public disclosure appear to have been significantly lower. This likely results from the decision to move toward the use of social media for public disclosure as opposed to more costly, traditional forms of media (e.g., newspaper and radio advertisements). This is consistent with a growing body of literature on the effectiveness of a social media campaign as a method for public disclosure in research.(9,18,20–25)

Some experts have expressed concerns that the cost and complexity of community consultation and public disclosure have slowed the conduct of randomized trials in emergency care(26). FDA guidance does suggest, however, that “IRBs may consider more or less community consultation necessary, depending on the unique circumstances of a particular study and the community,” giving the example that “less extensive community consultation may be appropriate for . . . . a study comparing an available treatment . . . with another product about which much is known.”(27) The relatively cost-efficient completion of community consultation and public disclosure in the RSI trial represents an attempt by trial leadership to follow this guidance, matching the extent of community consultation to the risks and burdens of participating in a trial comparing standard-of-care treatments. Ketamine and etomidate are the two most commonly used sedative medications during emergency tracheal intubation in many settings in the United States, both have been received by millions of patients each year for decades, the risk profiles of both medications are well known, and both medications are approved by the FDA for the studied indication at the studied doses. While the extent of community consultation and public disclosure in RSI were significant, reaching more than 1.2 million adults, they may not have been as intensive as prior trials evaluating novel, experimental, unapproved drugs or devices with potentially greater risks.

We also found that the costs of community consultation and public disclosure were driven largely by personnel costs rather than by fixed costs. Completion of community consultation and public disclosure required an average of 139 hours of research coordinator and physician site investigator time per site; a total of 833 hours across all sites. The time required varied significantly across sites, ranging from 63 hours to 261 hours. Several factors may explain this variability, including: site experience with community consultation and public disclosure processes, availability of non-study team local support (e.g. press office, local community engagement teams), variable reliance on the central engagement coordinator based at the coordinating center, and community-specific differences in acceptability of the studied interventions or the conduct of trials without prospective informed consent. Because the hours devoted to each activity were collected retrospectively, recall bias may also have contributed to the variability reported across sites.

Previous trials have taken different approaches to coordinating the completion of community consultation and public disclosure activities at participating sites, including: (i) a central conduct model(10) (site activities conducted centrally by the coordinating center), (ii) a federated model(19) (in which plans are designed centrally with efforts to oversee and align approaches while allowing local autonomy in planning and conduct), and (iii) a fully local model(28) (in which all sites generate plans without significant guidance or oversight from the coordinating center). The RSI trial used a federated model in which sites worked with their local community and IRB to develop a plan for community consultation and public disclosure using standardized templates developed by the coordinating center, which included required activities and a menu of optional activities and associated minimum standards. We found this approach to be successful in identifying and addressing community-specific concerns while facilitating efficient completion. All community consultation and public disclosure activities during the PCORI-funded multi-center stage of the trial were completed in nine months. The success of this model was greatly facilitated by having an experienced, central engagement coordinator to develop templates, identify community members and work with sites with less experience with EFIC trials.(1,9)

## LIMITATIONS

This article has several limitations. First, the pre-existing patient and community engagement infrastructure available at some participating sites in the RSI trial likely improved efficiency and decreased cost compared to sites that do not have pre-existing infrastructure, such as many non-academic hospital systems. Second, community consultation and public disclosure activities vary greatly in cost and effort, and decisions regarding which activities are chosen for a given trial may be related to the trial’s specific funding mechanism and budget.

Third, the extent of the patient and community engagement during trial conduct (not a requirement of EFIC) is based, in part, on recommendations regarding engagement provided by the funder, PCORI, and the resources required to conduct this level of engagement may not be available in every EFIC trial. Fourth, the time and cost of community consultation and public disclosure activities were estimated retrospectively, after completion of all activities, and recall bias could have resulted in underestimation of the total effort and cost. Fifth, estimates do not account for substantial effort from (i) non-research personnel at sites (e.g., media office personnel); (ii) coordinating center personnel (e.g., trial primary investigator, project managers) who led the design of templates, regulatory approval, oversite of site execution of activities, and submission of completed activities to the IRB and FDA; and (iii) the engagement coordinator who led the recruitment and preparation for local community engagement studios and town halls at sites that did not manage these locally.

## CONCLUSIONS

In preparation for conducting the RSI trial, study personnel completed community consultation that included bidirectional interactions with over 1,000 patients, families, and community members and public disclosure that reached over 1.2 million people living in the communities in which the trial is being conducted. The use of digital channels of communication for community consultation and public disclosure and an approach that matched the intensity of engagement activities to the risk of the research resulted in a total cost and duration of activities that were lower than reported in prior EFIC trials. However, these costs remained significant and may inform which treatments are feasible to examine in trials with EFIC.

## Data Availability

All data produced in the present study are available upon reasonable request to the authors.

## CONTRIBUTORS

All study authors approved the final version of this manuscript.

All study authors critically reviewed the manuscript for important intellectual content.

Study concept and design: MWS, JDC, AAL

Acquisition of data: All authors

Analysis and interpretation of data: AAL, TLI, SCD, MWS, JDC

Drafting of the manuscript: AAL, TLI, MWS, JDC

Revision of the manuscript: All authors

Acquisition of funding: MWS, JDC

## FUNDING

Research reported in this article was funded through a Patient-Centered Outcomes Research Institute (PCORI) Award (BPS-2022C3-30021). The views and statements presented in this article are solely the responsibility of the authors and do not necessarily represent the views of PCORI. Funding for the trial was also provided by the National Heart, Blood, and Lung Institute (NHLBI) (K23HL153584). The views and statements presented in this article are solely the responsibility of the authors and do not necessarily represent the views of the NHLBI.

## COMPETING INTERESTS AND FINANCIAL DISCLOSURES

The authors have no financial conflicts of interest relevant to the current work.

B.E.D. reported having received compensation from UpToDate unrelated to the current work.

A.A.G. reported having received compensation from Biomeme, SeaStar, and National Institutes of Health, unrelated to the current work.

S.A.T. reported having received compensation from the American College of Emergency Physicians unrelated to the current work.

D. W. R. reported having received compensation from ABSS Solutions, Quantum Leap Healthcare Collaborative, Critical Care Clinical Trial Network, Achieve Life Sciences, and Direct Biologics unrelated to the current work.

J.A.P. reported having received compensation from American College of Chest Physicians unrelated to the current work.

J.P.G. reported having received compensation from American College of Chest Physicians unrelated to the current work.

T.W.R. reported having received compensation from Cumberland Pharmaceuticals, Cytovale, Nestle, and Sanofi unrelated to the current work.

M.W.S. reported having received compensation from Baxter Healthcare Corporation, DynaMedex, and Reprieve Cardiovascular, Inc unrelated to the current work.

J.D.C. reported compensation from Reprieve Cardiovascular, Inc unrelated to the current work.

## Notes

### Author Declarations

The institutional review board (IRB) at Vanderbilt University Medical Center gave ethical approval for the RSI trial protocol (IRB number: 210500). The RSI trial protocol was also approved by the US Food and Drug Administration (FDA) (IND 141424). The study is being conducted with Exception from Informed Consent Requirements for Emergency Research (EFIC) (21 CFR 50.24).36 Plans for community consultation and public disclosure were approved by the single IRB at Vanderbilt University Medical Center. The IRB of each participating site provided local context for the community consultation and public disclosure plan and activities.

### Summary of Updates

Revision to author affiliations and inclusion of supplemental file 1 with list of collaborators.

