## Supplement 1 for "Community Consultation and Public Disclosure for the Randomized Trial of Sedative Choice for Intubation"

**List of RSI Investigators**

Coordinating Center: *Clinical Coordinating Center (Vanderbilt University Medical Center, Nashville TN)* – Jonathan D. Casey, MD MSc* (Director, Coordinating Center); Matthew W. Semler, MD MSc* (Chair, Steering Committee); Ariel A. Lewis, MPH, BSN, RN*; Jin H. Han, MD, MSc*; Wesley H. Self, MD, MPH*; Todd W. Rice, MD, MSc*; Bradley D. Lloyd, RRT-ACCS*; Cheryl L. Gatto, PhD; Grace Van Winkle, MPH; Sydney Vidrine, MPH; Tiffany L. Israel, MSSW*; Madison E. White, RN; Margaret A. Hays, RN MSN*.  *Data Coordinating Center (Vanderbilt University Medical Center, Nashville TN)* – Brant Imhoff, MS; Li Wang, MS; Matthew S. Shotwell, PhD.

Patient Representatives: Aida Strom (Minneapolis, MN); Barbara Gould, MSW (Denver, CO)*; Eileen Rubin, (Northbrook, IL); Jasmine McIntosh (Birmingham, AL); Patrick Luther (Nashville, TN); Sherman Transou (Winston-Salem, NC).

Denver Health Medical Center: Stacy A. Trent, MD MPH*; Carolynn Lyle, PA-C MPH*; L. Jane Stewart, MD JD MPH*.

Hennepin County Medical Center: Brian E. Driver, MD*; Aaron E. Robinson, MD MPH*; Matthew E. Prekker, MD MPH*; Julianna Prohofsky, BS.

Wake Forest School of Medicine: Kevin W. Gibbs, MD*; Jessica A. Palakshappa, MD MS*; J. Maycee Cain, BS*; John P. Gaillard, MD*; Ashley Strahley, MPH; Julia Narendra, MPH.

University of Alabama at Birmingham Medical Center and Heersink School of Medicine: Derek W. Russell, MD*; Logan L. Beach, MD*; Micah R. Whitson, MD*; Sheetal Gandotra, MD; Donna S. Harris, RN; Dianne Freeman, BS RRT; David Page, MD; Sonya Hardy, MA; Necole Harris, BS; Peter Morris, MD.

University of Colorado School of Medicine: Adit A. Ginde, MD MPH*; Cori Withers, BS*; Amy E. Sullivan, BA*; Kristine Schauer, MBA RN; Barbara Gould*; Daniel Resnick-Ault, MD; David J. Douin, MD MSc; Jason C. Brainard, MD; Neil R. Aggarwal, MD MHSc; Carrie Higgins, BSN; Laura G. Murphy, BS.

Vanderbilt University Medical Center: Stephanie C. DeMasi, MD*; Kevin P. Seitz MD, MSc*; Elizabeth M. Frawley, BSN RN*; Margaret A. Hays, RN MSN*.

*Denotes members of the writing committee who are listed as authors on the manuscript, the remainder of the RSI investigators represent collaborators.
